# Seroprevalence of Hepatitis B Virus Infection and Associated Factors Among Blood Donors at a District Hospital in Ghana: A 2-Year Retrospective Study

**DOI:** 10.64898/2026.09.10.26362725

**Authors:** Samuel Kyeremeh Adjei, Prosper Adjei, Esther Obeng

**Author notes:** Correspondence: Samuel Kyeremeh Adjei.

## Abstract

**Background and Aims:** Hepatitis B virus (HBV) infection remains a significant global public health concern, particularly in developing countries, where it poses a major challenge to safe blood transfusion services. Blood donors represent an important subset of the population for assessing HBV prevalence, as transfusions are a key route for transmission. This study aimed to determine the seroprevalence of HBV infection among blood donors at Methodist Hospital, Wenchi, Ghana.

**Methods:** A retrospective analysis was conducted using data from the blood bank at Methodist Hospital, Wenchi from January 2022 to December 2023 and analyzed using SPSS version 25.

**Results:** The overall HBV seroprevalence among blood donors was 6.8%, with a higher infection rate among females (16.7%) compared to males (6.3%). The highest seroprevalence (9.4%) occurred in donors aged 30–39 years and voluntary donors showed a higher infection rate (8.2%) than non-voluntary donors (6.5%). Among blood groups, individuals with blood groups A and B exhibited the highest infection rates (8.5%). Factors such as age (AOR: 2.62, 95% CI: 1.10 – 5.32, p-value =0.023), gender (AOR: 2.10, 95% CI: 1.05 – 4.35, p-value= 0.004) and Rhesus status (AOR: 1.68, 95% CI: 1.08 – 3.35, p-value= 0.031) were significantly associated with HBV infection.

**Conclusion:** The study revealed a significant HBV seroprevalence of 6.8% among blood donors, classifying the region as medium-endemic per WHO guidelines. The findings underscore the urgent need for enhanced HBV screening, rigorous blood safety measures and targeted public health interventions including vaccination and education campaigns to mitigate the transmission risk.

## 1 Introduction

Blood transfusion is a vital therapeutic intervention that greatly improves patient survival, particularly in situations involving significant blood loss or anemia. Nonetheless, it poses a risk of transmitting infections such as the human immunodeficiency virus (HIV), hepatitis B virus (HBV), syphilis and hepatitis C virus (HCV) ^1,2^. This risk stems from the potential contamination of donated blood with infectious agents, leading to the transmission of these infections.

The HBV is recognized as the most infectious blood-borne virus owing to its high infectivity (i.e. up to 100 times more infectious than HIV), capacity to survive outside the human body for at least one week and multiple transmission routes. Body fluids such as blood, semen and vaginal secretions are the main vehicles for the transmission of the virus ^3^. Studies have shown that blood transfusion is the most prevalent mode of transmission of HBV in developing nations aside vertical transmission ^4^. Worldwide, an estimated 240 million individuals are affected by chronic HBV infection, leading to more than 300,000 complications associated with liver diseases, including cirrhosis and liver cancer, which account for approximately 68,600 fatalities ^5,6^. The classification of HBV prevalence indicates that rates above 8% are deemed high, those ranging from 2% to 7% are classified as medium and rates below 2% are regarded as low endemicity ^7^.

Annually, over 81 million units of blood are donated worldwide and blood transfusion remains an indispensable medical intervention with no suitable substitute, vital for the survival of millions of patients in need of transfusions ^7,8^. Blood-borne infections, particularly HBV, pose a considerable challenge to the provision of safe blood transfusions. This issue is especially pronounced in regions where HBV is endemic and where effective screening techniques are scarce ^9^. The transfusion of contaminated blood accounts for around 16 million new cases of HBV infections worldwide, with each unit of blood carrying a 1.0% risk of transmitting blood-borne pathogens ^10^. Furthermore, it is estimated that unsafe blood transfusions will result in around 45,000 new cases of HBV infections in Africa each year. Additionally, approximately 1.6 million blood units are discarded annually as a consequence of blood-borne infections, including HBV ^11^. Consequently, 12.5% of individuals receiving transfusions in sub-Saharan Africa face the risk of acquiring HBV infections^12^. HBV infections continue to be an overlooked issue in Africa, affecting more than 60 million individuals across the continent ^13^.

The occurrence of HBV infections among blood donors in Africa exhibits significant regional variation across the continent, with prevalence rates varying between 5% and 7% ^11^. Specifically, it is reported to be 4.1% in Nigeria ^14^, 5.6% in Kenya ^12^, 10.9% in Ethiopia^15^ and 2.0% in Eritrea ^16^. The main risk factors associated with HBV transmission include engaging in unprotected sexual activities with multiple partners, sharing needles or syringes for injection, utilizing non-sterilized medical instruments and coming into contact with contaminated items, such as inadequately sterilized medical, surgical and dental tools ^14,17^. The World Health Organization (WHO) advocates for the rigorous screening of all blood donations to detect transfusion-transmissible infections, such as HIV, HBV and HCV. This recommendation has been embraced by Ghana to ensure safe transfusions of blood and blood products^18^.

Despite the implementation of various initiatives such as widespread vaccination campaigns and early childhood immunization programs aimed at reducing the transmission of HBV, the infection remains highly prevalent, particularly in developing nations like Ghana ^19^. There is a paucity of data on the prevalence of HBV infection among blood donors in the Bono region of Ghana. This study aimed to provide significant insights into the seroprevalence of HBV infection and its associated factors among blood donors at a district hospital in the Bono Region of Ghana.

## 2 Materials and Methods

### 2.1 Study Design and Setting

This study conducted a retrospective analysis using donors’ record from the blood bank of Methodist Hospital laboratory from January 2022 to December 2023. It is a primary level facility with six different specialties and also serves as the principal referral facility for the Wenchi Municipality and its environs. Due to its high admission rates, the hospital conducts periodic blood donation campaigns to meet the demand for blood transfusions. The hospital laboratory utilizes an immunochromatographic test, a single-step procedure designed to detect the presence of hepatitis B surface antigen (HBsAg) (QUINGDAO HIGHTOP BIOTECH CO. LTD., CHINA) with a specificity of 99.84% and sensitivity of 99.90%.

### 2.2 Study Population

The study included both voluntary and non-voluntary blood donors who visited the hospital’s blood bank during the designated study period. Non-voluntary donors in this context refers specifically to replacement donors, as commercial donation is not permitted at our facility. Participants were prospective donors aged 16 to 60 years who successfully completed a medical history assessment, as well as pre-donation screening tests that evaluated hemoglobin levels, blood pressure, pulse and weight. Additionally, these individuals underwent screening for Hepatitis B.

### 2.3 Data Retrieval, Processing and Analysis

Archived data on both voluntary and non-voluntary blood donors during the stipulated study period was retrieved from the hospital’s blood bank using Microsoft excel 2021. The blood transfusion service clinical record sheets of the laboratory and screening registers were reviewed and relevant data for the study was collated. Data on behavioural risk factors were not available. Unique donor identification numbers along with donor demographic information were meticulously used to verify donor records and ensure the elimination of duplicate entries. After the data was cleaned and verified, same was exported to IBM Statistical Package for the Social Sciences (SPSS Inc., Chicago, USA) version 25 for analysis. Descriptive statistics were reported as frequencies and percentages. Bivariate and multivariate logistic regression analysis were employed to determine factors associated with hepatitis B infection among blood donors.

### 2.4 Ethical approval

Ethical approval and waiver of informed consent were obtained from the Research Ethics Committee of Methodist Hospital, Wenchi, Ghana (Ethics approval reference: MHW/INT/56/02).

## 3 Results

Over the course of the study period, a total of 3835 prospective donors were screened for HBsAg at the blood bank. Of these, 3520 were included in the subsequent analysis while 315 were excluded due to incomplete and illegible data. Out of the total donors, 3352 representing 95.2% were males and 168 (4.8%) were females. The mean age of donors was 28.9 years with standard deviation of 8.2 years and ranged from 15 to 59 years. For male donors, their age ranged from 15 to 59 years with mean age of 28.4 years and standard deviation of 8.1 years. However, for female donors, their age ranged from 17 to 56 years with mean age of 27.9 years and standard deviation of 7.3 years. For HBsAg positive donors, their age ranged from 20 to 55 years with mean age of 31.2 years and standard deviation of 7.2 years. For males infected with HBV, their ages ranged from 20 to 55 years with mean age of 32.7 years and standard deviation of 9.2 years. On the other hand, the age range of infected females was 22 to 50 years with mean age and standard deviation of 30.5 years and 8.4 years respectively. Majority (55.7%, n=1961) of the donors were within 20 – 29 age bracket while 24.5% were within 30 – 39 age group [Table 1 and 2].

**Table 1:**
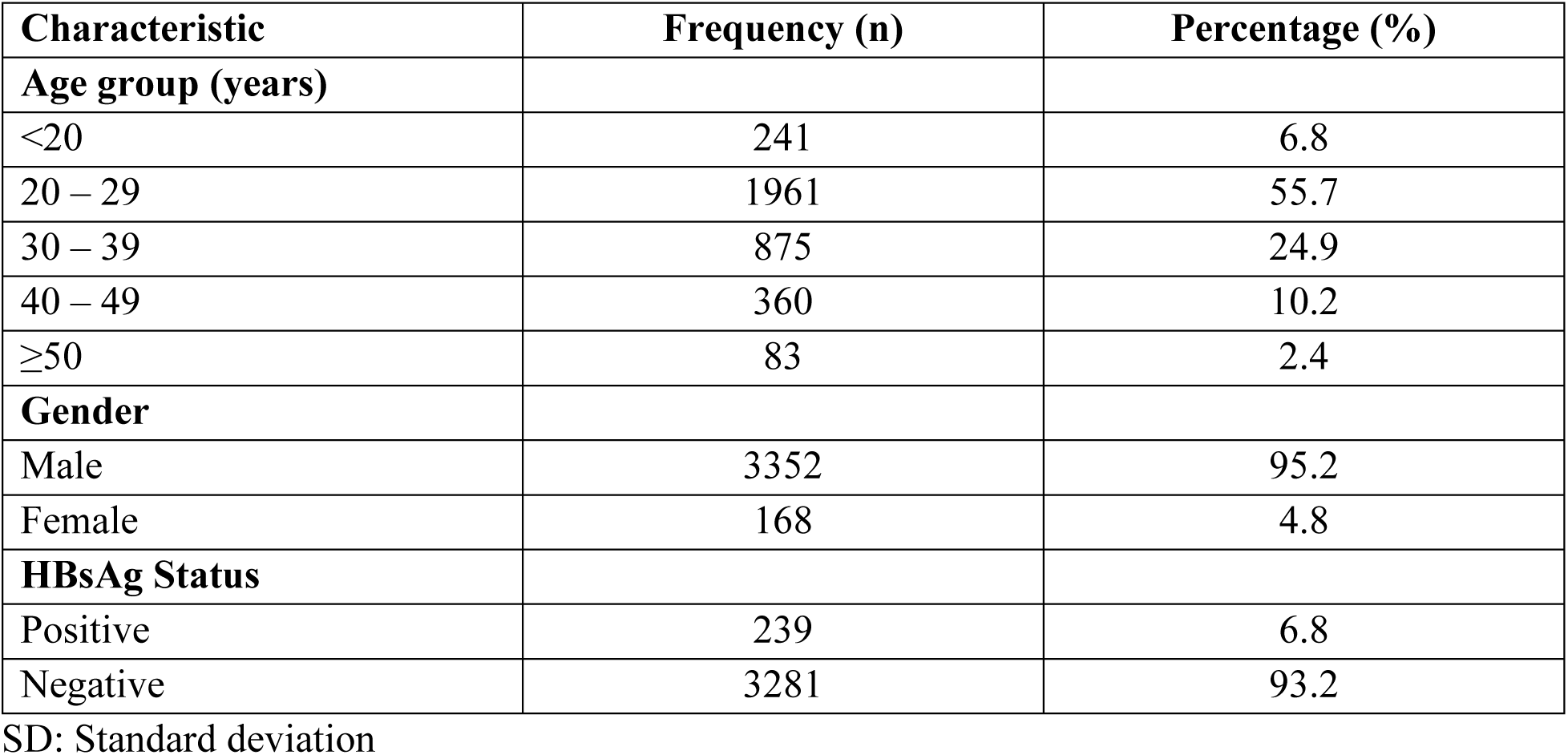
Demographic characteristics of blood donors at Methodist Hospital, Wenchi, Ghana (n=3520).

| Characteristic | Frequency (n) | Percentage (%) |
| --- | --- | --- |
| **Age group (years)** |  |  |
| <20 | 241 | 6.8 |
| 20 – 29 | 1961 | 55.7 |
| 30 – 39 | 875 | 24.9 |
| 40 – 49 | 360 | 10.2 |
| ≥50 | 83 | 2.4 |
| **Gender** |  |  |
| Male | 3352 | 95.2 |
| Female | 168 | 4.8 |
| **HBsAg Status** |  |  |
| Positive | 239 | 6.8 |
| Negative | 3281 | 93.2 |

SD: Standard deviation

**Table 2:** Age distribution and mean age of blood donors and HBsAg-positive individuals.

| Population | Age range | Mean age | Standard deviation |
| --- | --- | --- | --- |
| **Blood donors** | 15 – 59 | 28.9 | 8.2 |
| Male | 15 – 59 | 28.4 | 8.1 |
| Female | 17 – 56 | 27.9 | 7.3 |
| **HBsAg-positivity** | 20 – 55 | 31.2 | 7.2 |
| Male | 20 – 55 | 32.7 | 9.2 |
| Female | 22 – 50 | 30.5 | 8.4 |

HBsAg: Hepatitis B surface Antigen

A significant majority (83.1%, n=2925) of donors were non-voluntary donors while the remaining (16.9%, n= 595) were voluntary donors. In the present study, 239 donors tested positive for HBsAg representing an overall seroprevalence of 6.8%. Of the 2925 non-voluntary donors, 6.5% were seropositive for HBsAg while 8.2% were seropositive among voluntary donors. With regard to the study period, the seroprevalence of HBsAg among donors was 6.7% in 2022 and 6.8% in the year 2023. The highest infection rate was found among female donors (16.7%), donors aged 30 – 39 years (9.4%) and among voluntary donors (8.2%). The annual analysis revealed an increase in HBV infections within the study population over the observed period [Table 3].

**Table 3:**
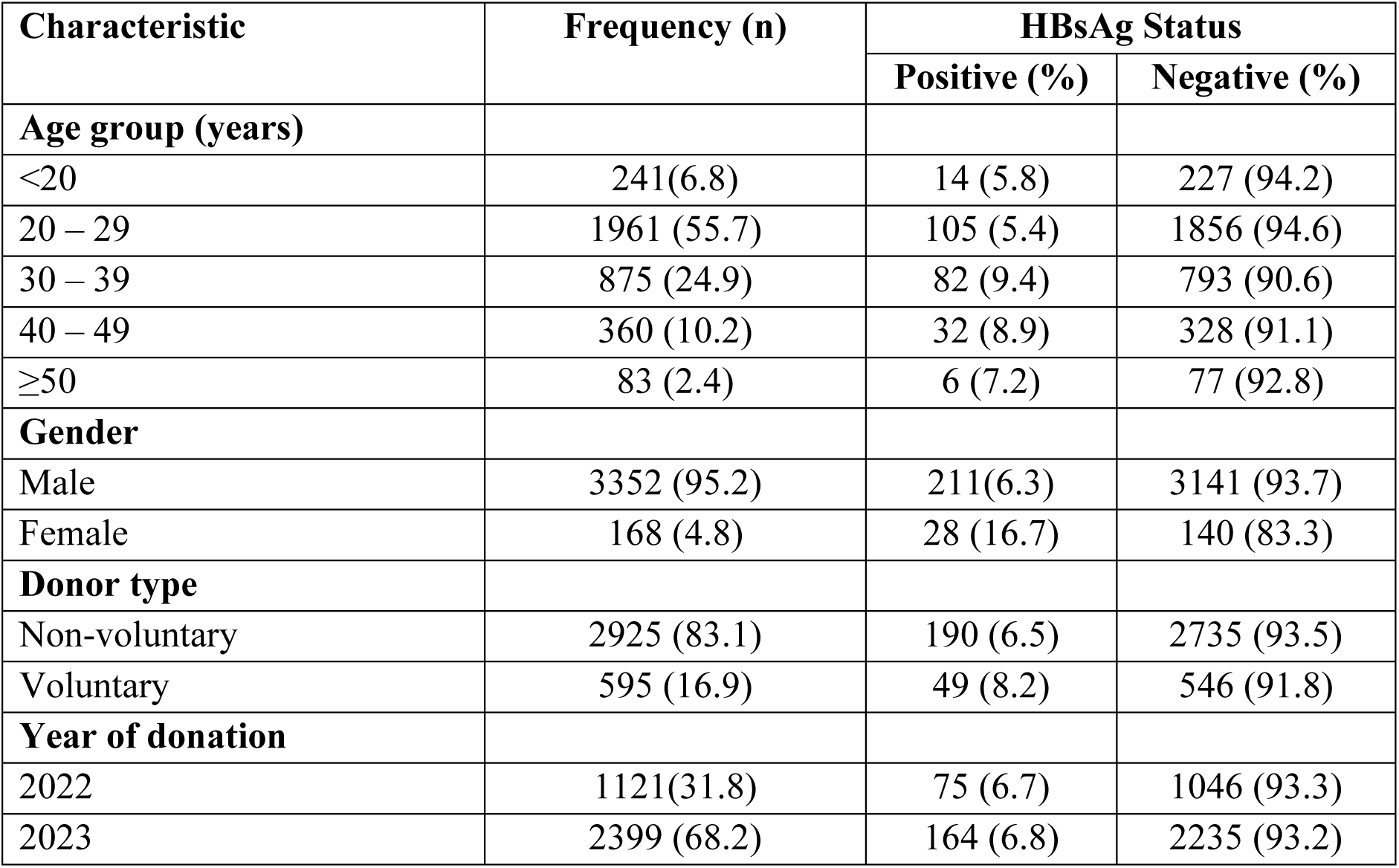
Prevalence of hepatitis B infection stratified by age group, gender, donor type and year of donation (n=3520).

The predominant blood group among the donors was found to be group O, accounting for 55.8% of the total, followed by blood group B at 24.2%. Blood groups A and AB represented 19.3% and 0.7% respectively. Regarding Rhesus (RH) factor, 91.5% of the donors were found to be Rh-D positive, whereas the remaining 8.5% were Rh-D negative. With regard to the prevalence of HBV infection among ABO blood groups and Rh-D status of donors, the highest rate of infection was found among donors with blood groups A and B (8.5%), followed by group O (5.4%), while the lowest rate was recorded among blood group AB. Individuals with Rh-D negative blood type demonstrated a higher infection rate (7.7%) compared to those with the Rh-D positive genotype, who experienced an infection rate of 6.7% [Table 4].

**Table 4:** Distribution of hepatitis B infection among the ABO blood group and Rhesus status of donors (n=3520).

| Characteristic | Frequency (n) | HBsAg Status |  |
| --- | --- | --- | --- |
|  |  | Positive (%) | Negative (%) |
| **Blood group** |  |  |  |
| A | 679 (19.3) | 58 (8.5) | 621(91.5) |
| B | 852 (24.2) | 72 (8.5) | 780 (91.5) |
| O | 1964 (55.8) | 107 (5.4) | 1857 (94.6) |
| AB | 25 (0.7) | 2 (8.0) | 23 (92.0) |
| **Rhesus status** |  |  |  |
| Positive | 3221 (91.5) | 216 (6.7) | 3005 (93.3) |
| Negative | 299 (8.5) | 23 (7.7) | 276 (92.3) |

The study revealed that factors such as age (AOR: 2.62, 95% CI: 1.10 – 5.32, P =0.023), gender (AOR: 2.10, 95% CI: 1.05 – 4.35, P= 0.004) and Rhesus status (AOR: 1.68, 95% CI: 1.08 – 3.35, P= 0.031) were significantly associated with hepatitis B infection among blood donors [Table 5].

**Table 5:** Bivariate and multivariate logistic regression analysis of factors associated with hepatitis B virus infection among blood donors at Methodist Hospital, Wenchi, Ghana.

| Characteristic | HBsAg Status |  | Bivariate Analysis |  | Multivariate Analysis |  |
| --- | --- | --- | --- | --- | --- | --- |
|  | Positive<br>(n=239) | Negative<br>(n=3281) | COR (95% CI) | P-value | AOR (95% CI) | P-value |
| **Age group** |  |  |  |  |  |  |
| <20 | 14 (5.8) | 227 (94.2) | 1 |  | 1 |  |
| 20 – 29 | 105 (5.4) | 1856 (94.6) | 1.20 (0.91 – 1.56) | 0.021 | 1.36 (0.501 – 3.38) | 0.188 |
| 30 – 39 | 82 (9.4) | 793 (90.6) | 1.81 (1.14 – 3.21) | 0.015 | 2.62 (1.10 – 5.32) | 0.023* |
| 40 – 49 | 32 (8.9) | 328 (91.1) | 1.30 (0.57 – 4.41) | 0.134 | 1.03 (0.62 – 1.94) | 0.342 |
| ≥50 | 6 (7.2) | 77 (92.8) | 0.82 (0.51 – 2.10) | 0.627 | 0.99 (0.23 – 1.46) | 0.719 |
| **Gender** |  |  |  |  |  |  |
| Female | 28 (16.7) | 140 (83.3) | 1 |  | 1 |  |
| Male | 211(6.3) | 3141 (93.7) | 1.42 (0.73 – 3.12) | 0.023 | 2.10 (1.05 – 4.35) | 0.004* |
| **Donor type** |  |  |  |  |  |  |
| Voluntary | 49 (8.2) | 546 (91.8) | 1 |  | 1 |  |
| Non-voluntary | 190 (6.5) | 2735 (93.5) | 1.26 (0.93 – 1.98) | 0.003 | 3.18 (1.20 – 6.05) | 0.013* |
| **Blood group** |  |  |  |  |  |  |
| A | 58 (8.5) | 621(91.5) | 1 |  | 1 |  |
| B | 72 (8.5) | 780 (91.5) | 0.88 (0.213 – 2.01) | 0.931 | 1.10 (0.30 – 1.22) | 0.286 |
| O | 107 (5.4) | 1857 (94.6) | 0.75 (0.22 – 1.64) | 0.335 | 0.91 (0.22 – 1.50) | 0.503 |
| AB | 2 (8.0) | 23 (92.0) | 0.89 (0.47 – 1.39) | 0.212 | 0.78 (0.15 – 1.28) | 0.604 |
| **Rhesus status** |  |  |  |  |  |  |
| Positive | 216 (6.7) | 3005 (93.3) | 1 |  | 1 |  |
| Negative | 23 (7.7) | 276 (92.3) | 1.15 (0.51 – 3.20) | 0.018 | 1.68 (1.08 – 3.35) | 0.031* |

\*: Significant, COR: Crude odds ratio, AOR: Adjusted odds ratio

## 4 Discussion

Blood transfusion plays a crucial role in healthcare, with millions of lives saved worldwide annually through this vital procedure ^20^. In Ghana, numerous individuals frequent the blood banks of different hospitals and blood donation centers nationwide to donate blood for individuals in need of transfusions for survival. This study revealed a preponderance of non-voluntary donors, male donors and individuals aged 30 years or younger. These findings align with earlier studies carried out in various regions of Ghana ^21,22^. Comparable findings were also documented in Nigeria and India ^23,24^. While definitive explanations for the consistent prevalence of male dominance in the donor population among Ghanaians remain incomplete, it can be inferred that various factors including specific beliefs, socio-cultural practices and the tendency of males to take initiative in decision-making over females may contribute to this ongoing trend ^20,21^. One prominent explanation for the disparity in blood donation rates between genders is that menstruation and childbirth significantly limit the number of female donors. Individuals in these categories are often prohibited or deferred from participating in blood donation. Additionally, there exists a widespread misconception that blood donation is primarily a male activity, which further discourages female participation ^25^. Insufficient awareness regarding the importance of voluntary blood donation may contribute to the low participation rates among voluntary donors. Conversely, the prevalence of younger donors in this study may be attributed to a growing sense of social responsibility among younger individuals as well as influence by their peers and the social environment in which they operate.

A total of 239 donors tested positive for HBsAg, resulting in a sero-positivity rate of 6.8% which aligns with the rates reported in earlier studies ^26,27^. Other studies conducted earlier recorded relatively higher prevalence of HBsAg seropositivity ^28,29^. The findings of the present study together with those previously mentioned, confirm Ghana as an endemic region for HBV. Insufficient awareness and the residents’ inability to bear the expenses associated with receiving the three doses of the hepatitis B vaccine could account for the high prevalence of HBV infection in Wenchi and other parts of Ghana.

In line with worldwide trend, males constituted the predominant group within both the donor population and the instances of HBV infection. Nonetheless, the seroprevalence of HBV among female donors was 16.7% compared to 6.3% among their male counterparts. This finding aligns with reports of earlier studies ^30,31,32^. This gender disparity may be attributed to factors such as increased biological vulnerability in females, cultural practices limiting access to healthcare and socio-economic barriers that hinder education on HBV prevention and screening. These findings underscore the need for gender-specific public health interventions to reduce HBV transmission. Gender-specific interventions may include targeted education programs for women, improving access to HBV screening and vaccination for females, incorporating HBV education into maternal and reproductive health services and addressing socio-cultural barriers that limit female access to healthcare. In contrast to the finding of this study, other studies recorded high prevalence among male donors ^33,34^. Although crude prevalence indicated that female donors had a higher HBV infection rate (16.7%) compared to males (6.3%), multivariate regression analysis of this study demonstrated that male donors were 2.10 times more likely to be infected with HBV compared to their female counterparts. This apparent discrepancy underscores the importance of distinguishing between descriptive prevalence and adjusted odds. The higher crude prevalence among females may reflect their small representation in the donor pool and clustering of risk factors such as age or donor type. Regression analysis, which controls for these confounders, identifies males as the independent high-risk group. Males are often more prone to engage in high-risk behaviours such as unprotected sex and intravenous drug use which increase the likelihood of HBV transmission. Additionally, males may be less likely to seek regular medical check-up leading to a high chance of HBV infection.

The highest prevalence of HBV among blood donors was found among individuals aged 30 – 39 years. This finding is similar to what was reported by earlier studies^22^. The multivariate regression analysis affirms that donors aged 30 – 39 years were 2.62 times more likely to be infected with HBV compared to their younger counterparts. This trend might reflect cumulative exposure to risk factors such as unsafe sexual practices, use of non-sterile medical equipment or lack of vaccination in earlier life stages, particularly for individuals born before the widespread implementation of childhood immunization programs^35^. In contrast, other studies reported high seroprevalance among donors aged 20 – 29 years ^20,21^.

The higher seroprevalence among voluntary donors (8.2%) compared to non-voluntary (replacement) donors (6.5%) is an intriguing observation. This finding agrees with the results of earlier studies^36^. Voluntary donors are often presumed to be at lower risk due to stricter pre-donation screening and self-selection. However, this finding may reflect a lack of awareness or education about HBV transmission among this group. The lack of awareness may also reflect gaps in the quality and standard of counselling and education available to voluntary donors. Alternatively, it could be indicative of the broader socio-demographic characteristics of voluntary donors in this setting, which may include individuals with limited access to preventive healthcare. In contrast, the regression analysis revealed that non-voluntary donors were 3.18 times more likely to be infected with HBV. This divergence highlights the role of confounding factors. Non-voluntary donors often donate under specific conditions or due to compulsion. They may not be fully informed about their health status or may lack adequate knowledge regarding the blood donation process.

Interestingly, the distribution of HBV infection among blood donors by blood group showed that individuals with blood groups A and B had the highest seroprevalence (8.5%), while those with blood group O exhibited a lower rate (5.4%). Similarly, Rh-D negative individuals demonstrated a higher seroprevalence (7.7%) compared to Rh-D positive individuals (6.7%). The study also found that Rhesus negative donors were 1.68 times more likely to be infected with HBV compared to Rhesus positive donors. While the biological significance of these associations remains unclear, these findings may warrant further investigation to determine if certain blood groups or Rhesus factor confers increased susceptibility to HBV. This could have implications for donor selection and transfusion safety in endemic regions.

The findings of this study have significant implications for clinical, public health and research interventions. From the clinical perspective, the relatively high prevalence of HBV infection among blood donors poses a significant challenge to ensuring safe blood transfusion practices. Although the Methodist Hospital employs immunochromatographic testing with high sensitivity and specificity, this study underscores the pressing need for more advanced and rigorous HBV screening methods such as nucleic acid testing (including HBV DNA Polymerase Chain Reaction) and anti-HBc (Hepatitis B core antibody) testing to further reduce the risk of transfusion-transmitted HBV infections. Enhanced screening protocols should be complemented by robust donor education programs to mitigate the risk of HBV transmission. Clinicians must also recognize the broader implications of high HBV prevalence among blood donors, as it reflects the burden of the disease within the general population. This necessitates the implementation of a two-pronged strategy: improving the safety of blood transfusion services while also strengthening community-level HBV prevention and control measures. Additionally, clinicians should be vigilant in counseling patients on the risks of HBV transmission and the importance of vaccination, particularly for high-risk groups.

From the public health stance, the findings of this study highlight the urgent need for national and regional health authorities to prioritize HBV prevention and control. While Ghana has made notable progress in implementing vaccination programs and adhering to WHO guidelines for blood donor screening, gaps remain in achieving universal HBV vaccination coverage and ensuring equitable access to preventive healthcare services. Policymakers should consider scaling up efforts to expand access to HBV vaccination, particularly for adults and high-risk populations who may have missed routine childhood immunizations. Subsidizing the cost of HBV vaccines could significantly reduce the burden of the disease. Additionally, public health campaigns should focus on raising awareness about HBV transmission, encouraging voluntary testing and addressing cultural and socio-economic barriers that hinder access to care. The study also underscores the importance of enhancing the infrastructure and resources available for blood transfusion services in Ghana. This includes investing in advanced screening technologies, improving donor recruitment and retention strategies and ensuring the availability of safe blood and blood products. Furthermore, policymakers should explore strategies to increase female participation in blood donation by addressing misconceptions and barriers related to gender roles and health practices.

From research standpoint, this study contributes valuable data to the limited body of knowledge on HBV seroprevalence among blood donors in the Bono region of Ghana. However, several areas warrant further investigation. Firstly, longitudinal studies are needed to explore the factors driving the observed trends in HBV prevalence, including the role of socio-cultural, economic and behavioral determinants. Secondly, research should focus on evaluating the effectiveness of current HBV prevention and control initiatives, such as vaccination campaigns and public education programs to identify areas for improvement. The observed associations between HBV seroprevalence and blood group or Rhesus factor warrant additional research to determine whether these correlations are incidental or indicative of underlying biological mechanisms. Similarly, studies investigating the genetic or immunological factors contributing to HBV susceptibility could provide valuable insights into disease prevention and management. Finally, future research should explore innovative strategies for improving blood transfusion safety in resource-limited settings. This includes evaluating the cost-effectiveness of advanced screening technologies, assessing the feasibility of implementing universal HBV vaccination for adults and identifying best practices for donor recruitment and retention.

Despite this study contributing significantly to the trends of hepatitis B infection among blood donors in Ghana, it presents some limitations. The study relied on archived data which may have limited the completeness and accuracy of the information collected. Additionally, the study was conducted at a single primary care hospital, potentially limiting the generalizability of the findings to other healthcare settings or populations in Ghana.

## 5 Conclusion

This study highlights the continued burden of HBV infection among blood donors in Ghana, with an overall seroprevalence of 6.8%. The findings emphasize the need for comprehensive public health measures, including more rigorous HBV screening protocols, expanded access to vaccination programs, and targeted education initiatives to reduce HBV transmission. The higher seroprevalence among female donors, voluntary donors, and individuals in the 30–39 age group suggests the need for tailored interventions to minimize HBV transmission in these high-risk groups. Additionally, the association between blood groups, Rhesus factor and HBV seroprevalence warrants further investigation to understand potential underlying biological mechanisms. To ensure safer blood transfusion practices, healthcare authorities should invest in advanced screening technologies and strengthen blood donation systems. These efforts, combined with robust public health strategies, are vital for reducing the burden of HBV and ensuring the availability of safe blood for transfusion in Ghana.

## Acknowledgement

The authors are very grateful to the Manager and staff of Methodist Hospital Laboratory Department, especially staff of the blood bank for their immense assistance during data collection.

## Data availability statement

The datasets used and/or analyzed during this study are available from the corresponding author upon reasonable request.

## Ethics statement

Ethical approval was obtained from the Research Ethics Committee of Methodist Hospital Wenchi before commencement of the study (Ethics approval reference: MHW/INT/56/02). The study did not incorporate any identifying details such as patient name or identification number, ensuring that all data was handled with the highest level of confidentiality. Additionally, the requirement for informed consent was waived by the Research Ethics Committee.

## Author contributions

**Samuel Kyeremeh Adjei**: Conceptualization; Data analysis; Writing-original draft, Writing review and editing, **Prosper Adjei**: Writing-review and editing, **Esther Obeng**: Data curation. All authors have read and approved the final version of the manuscript. The corresponding author has full access to all of the data in this study and takes complete responsibility for the integrity of the data and the accuracy of the data analysis.

## Abbreviations

HBV: Hepatitis B virus
HBsAg: Hepatitis B surface antigen
HCV: Hepatitis C virus
HIV: Human immunodeficiency virus
NAT: Nucleic acid testing
WHO: World Health Organization
RH-D: Rhesus D
SPSS: Statistical package for social science

